# Assessing Holistic Health Interventions and Approaches for Individuals with Alcohol Use Disorder (AUD): Protocol for a Scoping Review

**DOI:** 10.1101/2025.06.04.25328277

**Authors:** Shubhi Nanda, Anna Roberts, Gisela Butera, Donna Barnett, Gwenyth R. Wallen, Jennifer J. Barb

## Abstract

**Introduction:** Alcohol Use Disorder (AUD) is a complex condition that requires a multidisciplinary approach to treatment. A comprehensive plan must address the physical, emotional, and mental health of the individual, which is where holistic health comes into play. Holistic health is a system of wellness that considers the whole person—body, mind, and spirit—and aims to restore balance and well-being through the combination of physical, mental, and emotional care. This approach typically incorporates natural and alternative forms of treatment along with conventional medical treatment. In recent years, there has been growing interest in integrating holistic health strategies into addiction treatment. The primary aim of this scoping review is to identify and characterize holistic health interventions currently implemented in the treatment for individuals with AUD. A secondary aim of this review is to collate the efficacy of the interventions as a treatment modality for people with AUD. Additionally, this review will identify gaps in the literature and suggest potential areas for future research.

**Methods and Analysis:** The review will adhere to the Arksey and O’Malley methodological guidelines for scoping reviews. Published research and pilot clinical trials that report on the utilization of holistic health interventions in AUD will be the focus of the search strategy. A total of 5 databases were searched: PubMed/MEDLINE, Embase, CINAHL, Web of Science, and Scopus. After screening, data will be extracted and synthesized from the included studies according to the guidelines developed in this protocol.

There will be no date restrictions used in the search. There is a language restriction of only articles published in English. To be included in the study, at least one type of a holistic intervention treatment (e.g., meditation, homeopathy, special diets, etc.) must have been employed. Excluded from the review will be any population that does not have hazardous drinking behaviors as associated with AUD, as well as any pharmaceutical interventions that are not coupled with a holistic intervention. The review itself will be an iterative process completed by two reviewers, and tie breakers will be done through discussions between the two reviewers to come to a unanimous consensus. Documents will be undergoing a risk assessment of bias for the authors to comment on the quality of the literature in this area. Finally, results will be discussed with all coauthors.

**Ethics and Dissemination:** This review will provide an overview of holistic therapies and interventions used to treat AUD, aiming to clarify their roles and effectiveness to support recovery. This review seeks to identify gaps in the literature and will recommend areas for additional research by identifying the strategies currently in use. The results of this review will be published in a peer-reviewed journal and made accessible to a general audience. No ethical approval is needed because this is a scoping review of the material that is already accessible.

**Article Summary:** *Strengths and limitations of this study:*

- A framework is outlined to extract information for holistic health interventions used for the treatment of alcohol use disorder while assessing the efficacy of the approaches compiled from this review. This review will also collate treatments that are used in concert with pharmacological intervention approaches for the treatment of AUD.
- This scoping review will be conducted using established methodological frameworks, including the Arksey and O’Malley framework and the PRISMA-ScR (Preferred Reporting Items for Systematic Reviews and Meta-Analyses Extension for Scoping Reviews) checklist, to ensure transparency and reproducibility of the process.
- The aim is to synthesize a wide range of interventions using diverse Medical Subject Headings (MeSH) to better characterize the development of these interventions in a heterogeneous population.
- A standardized data extraction form will be used to systematically capture and synthesize relevant study characteristics and outcomes across sources, supporting structured summarization by the review team.
- The reviewers represent a range of perspectives and academic backgrounds, which will help to address any bias in terms of the interpretation of the reviewed studies.
- Due to resource and reviewer language constraints, only English-language articles will be included, which may limit the generalizability and global applicability of the review findings.

## Introduction

### Background

Alcohol Use Disorder (AUD) is identified by the Diagnostic and Statistical Manual of Mental Disorders (DSM) characterized by the excessive intake of alcohol, resulting in considerable suffering in an individual’s social, personal, and professional lives (1). AUD is prevalent in the United States, contributing to public health concerns like accidents, violence, and long-term health problems which places a significant economic burden on the healthcare system (1, 2). Traditional treatment for AUD most commonly focuses on pharmaceutical interventions, but there is growing interest in incorporating holistic health approaches, which emphasize the interconnectedness of an individual’s mental, physical, and emotional well-being (3, 4). These approaches may complement or offer alternative benefits to traditional methods, potentially enhancing recovery outcomes in AUD populations (5).

Holistic health focuses on treating the whole person, not just their symptoms, by fostering balance across various aspects of life. This approach may incorporate practices such as mindfulness, nutrition therapy, exercise, psychotherapy, acupuncture, yoga, meditation, breathing exercises, herbal remedies, social support, spirituality/religion, and stress management techniques. By balancing the physical, emotional, and mental aspects of an individual’s well-being, holistic health seeks to address the root causes of addiction while promoting long-term healing. Cognitive-behavioral therapy (CBT) and motivational interviewing (MI), both of which focus on refocusing thoughts, behaviors, and motivations, can also be seen as part of this holistic approach since they integrate the psychological and emotional aspects of the individual into treatment, fostering healthier coping mechanisms (6). Despite growing interest in these practices, the research on how holistic interventions can be integrated into AUD treatment remains limited, with much of the focus still on pharmaceutical solutions like medication-assisted treatment (MAT) and the use of medications like disulfiram or naltrexone to manage cravings (7, 8).

### Study Rationale

A significant gap exists in the literature regarding the exploration of holistic practices in the treatment of AUD. While pharmaceutical interventions can effectively manage cravings and withdrawal symptoms, they often fall short in addressing the root causes of addiction and the individual’s overall well-being, factors critical for sustained recovery(4). In contrast, holistic approaches emphasize the treatment of underlying contributors such as chronic stress, unresolved trauma, mental health conditions, and lifestyle imbalances that may predispose individuals to AUD(9, 10).

Despite the demonstrated efficacy of behavioral therapies such as CBT and MI, limited research explores their integration with complementary holistic interventions, including group therapy, mindfulness-based practices, or nutritional support. These combined approaches may enhance outcomes by mitigating withdrawal effects and promoting long-term abstinence (6). Additionally, socio-cultural influences—such as family dynamics and cultural norms surrounding alcohol use, are pivotal in both the development and recovery from AUD, yet they remain underrepresented in conventional treatment paradigm(11).

Some studies have explored the intersection of holistic health interventions and AUD treatment, like nutritional therapies. For example, a 2024 study, exploring the effects of dietary intake, examined the impact of fiber intake on the gut microbiome of individuals with AUD, where it was found the a fiber rich diet helped to curb alcohol cravings during withdrawal and can help with staying abstinent (10). This research suggests that diet may influence mood and alcohol cravings, highlighting the potential for nutritional interventions as part of a holistic treatment plan (12). However, comprehensive reviews of the available literature on this topic are scarce, presenting an opportunity for further exploration.

### Study Objectives

This review aims to fill a critical gap in the literature by synthesizing existing research on the use of holistic practices in the treatment of AUD. Specifically, it will examine a range of treatment modalities, including mind-body interventions, nutritional therapies, and psychosocial approaches, to provide a more comprehensive perspective on AUD recovery. The review will also explore how these holistic strategies can be integrated with conventional treatments, with the goal of enhancing both immediate recovery outcomes and long-term well-being. Ultimately, this work seeks to advance understanding of the role holistic health practices may play in supporting sustained recovery from AUD.

## Methods and Analysis

### Protocol Design

A scoping review will be conducted to provide an overview of the presence of holistic interventions, their characteristics, and key factors of implementation as reported in the supporting literature. The review will follow the methodological framework developed by Arksey and O’Malley and updated by Joanna Briggs Institute (JBI) methodology for scoping reviews and the Preferred Reporting Items for Systematic Reviews and Meta-Analyses extension for Scoping Reviews (PRISMA-ScR) guidelines (13-15). The five stages of the methodological framework for scoping reviews as developed by Arksey and O’Malley (13, 16) will be used.

### Stage 1: Identifying the research question

The overarching aim of this review is to synthesize existing research on the use of holistic practices in the treatment of AUD as well as highlight corresponding gaps in the literature. The research team defined the following three research questions:

1. What holistic interventions are currently being implemented in the treatment of AUD and how do these interventions impact AUD recovery outcomes?
2. Which holistic interventions have been effective in treating AUD?
3. What are the perceived barriers to implementing holistic approaches in AUD treatment?
4. What are the common themes and outcomes reported in qualitative studies on holistic health and AUD?

A search of the following search engines will be conducted in the following databases:

#### Databases searched

- PubMed/MEDLINE (National Library of Medicine)
- Embase (Elsevier)
- CINAHL Plus (EBSCOhost
- Web of Science Core Collection (Clarivate Analytics)
- Scopus (Elsevier)

### Stage 2: Identifying relevant studies

To develop our search strategy, the three-step approach outlined in the JBI Scoping Review Guidance will be applied (17). The search strategy will be developed using relevant Medical Subject Headings (MeSH) terms and additional keywords related to AUD and various holistic health interventions. For a full list of search terms, see Supplemental Table 1. The search strategy will be created in collaboration with the NIH Library to ensure alignment with the objectives of this scoping review. Searches will be undertaken from database inception, with no restrictions on publication date or language. All references retrieved from the searches will be exported to Clarivate EndNote version 21, where duplicates will be identified and removed. The search strategy aims to locate both published and unpublished studies. We will include primary studies using any methodology (including experimental, quasi-experimental, observational, mixed-method, and qualitative study designs), which explore the use of holistic health interventions in treating AUD. Case reports, editorials, letters, or opinion pieces will not be included. Additionally, we will perform backward and forward searching of the reference lists of included papers to identify any additional relevant studies.

**Table 1:**
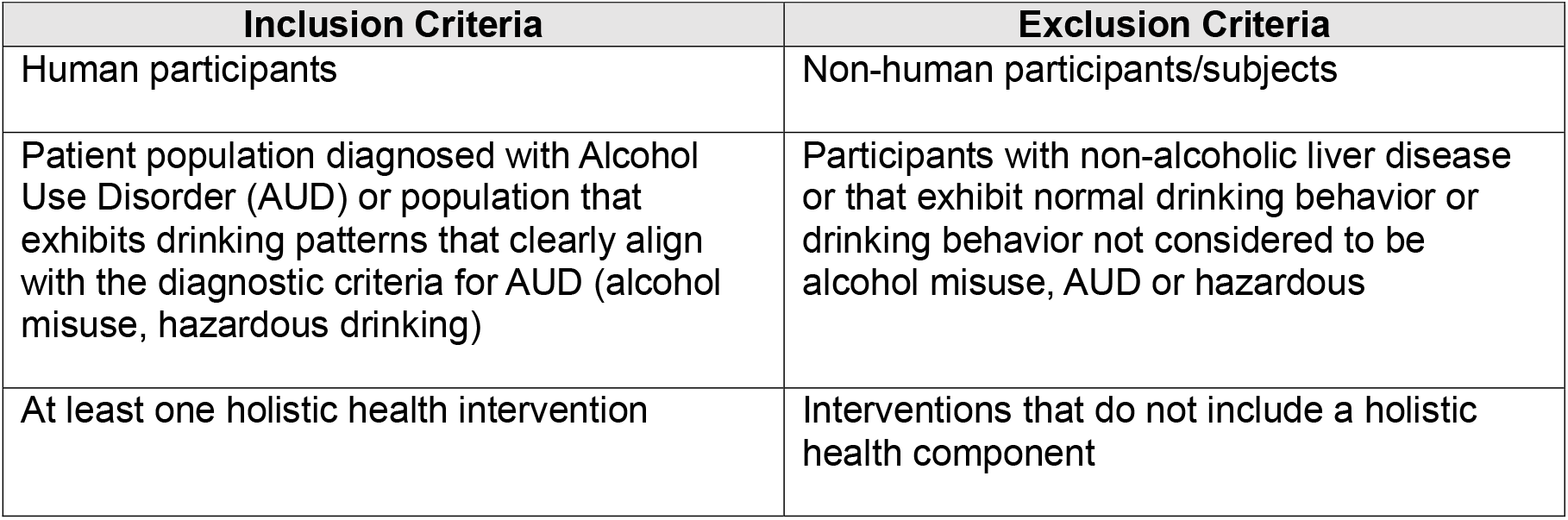
Inclusion and Exclusion Criteria for determining review selection.

| Inclusion Criteria | Exclusion Criteria |
| --- | --- |
| Human participants | Non-human participants/subjects |
| Patient population diagnosed with Alcohol Use Disorder (AUD) or population that exhibits drinking patterns that clearly align with the diagnostic criteria for AUD (alcohol misuse, hazardous drinking) | Participants with non-alcoholic liver disease or that exhibit normal drinking behavior or drinking behavior not considered to be alcohol misuse, AUD or hazardous |
| At least one holistic health intervention | Interventions that do not include a holistic health component |

### Stage 3: Study selection

Following the database searches, all identified citations will be transferred to Covidence, a screening software, and additional duplicates will be excluded (18). The titles and abstracts will be independently screened against the eligibility criteria by two researchers (SN and AR) (Table 1). The full texts of all potentially eligible articles will be retrieved, read in full, and assessed against the eligibility criteria. The title/abstract screen and full-text assessment will be completed by two to three members of the review team. Any discrepancies regarding the eligibility of an article at any stage will be resolved through discussion, and if necessary, an additional reviewer will be involved. If eligibility remains unclear, these discussions can be extended to the full review team. We will produce a Preferred Reporting Items for Systematic Reviews and Meta-Analyses (PRISMA) flowchart of the study inclusion process, recording the number of articles retained at each screening stage and the reasons for exclusion at the full-text screening stage (15). Covidence will be used to store and organize all articles. We will also record, tabulate, and report papers not available in English but which otherwise meet our eligibility criteria.

### Stage 4: Charting the data

Prior to the data extraction phase, the team will need to confirm access to all publications and gain access to publications that are unavailable through the National Institutes of Health (NIH) Inter-library loan system if possible. If the full text of the original paper is unavailable, then the paper will be excluded from the review and reported as such. In addition, we will contact corresponding authors of included articles for information that is either missing or unclear.

Through Covidence, a custom template for data extraction will be used to extract the data as they appear in the full-text review. Inter-rater reliability will be assessed through a two-reviewer method where they will compare the extracted data; if there is agreement between the two reviewers for the papers and the data extracted, then the studies will be divided amongst all reviewers. If there are any issues with the review process, then the team can meet to discuss and reevaluate the process to ensure reliability.

The following information will be charted as part of this scoping review:

### Stage 5: Collating, summarizing, and reporting the results

The holistic strategies found will be compiled and presented from the included research to determine the current approaches to managing and treating AUD (Table 2). Each intervention’s description, goal, and frequency of usage will be included in this table. Additionally, we will look at and report on any assessment or outcome measures that are used to gauge how beneficial these interventions are, as well as how frequently they are employed in different research.

**Table 2:**
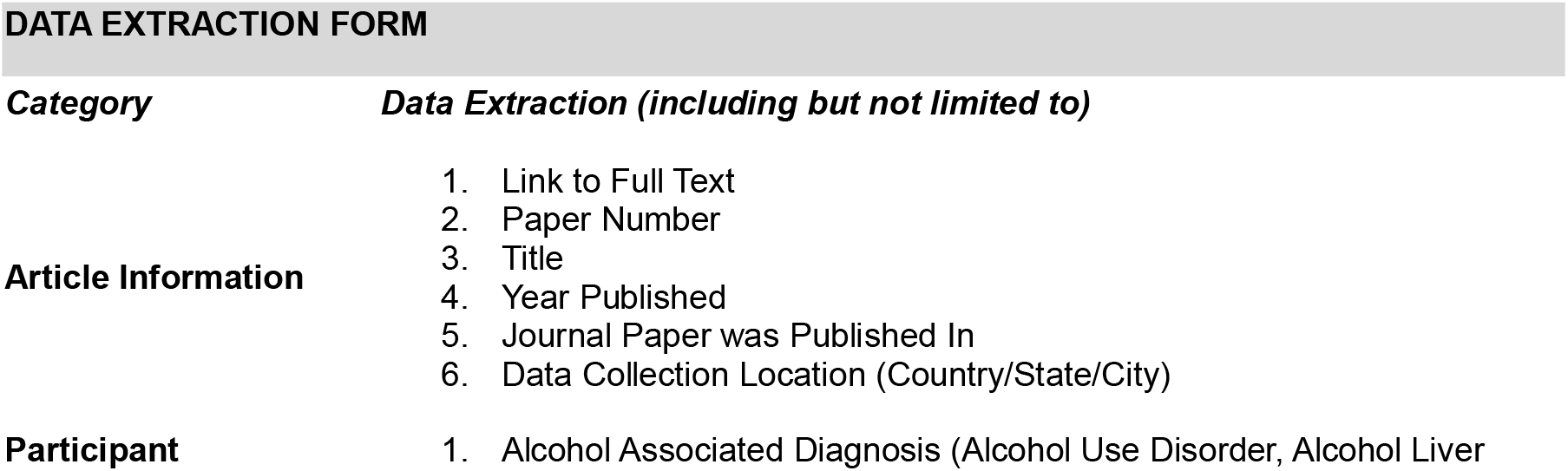
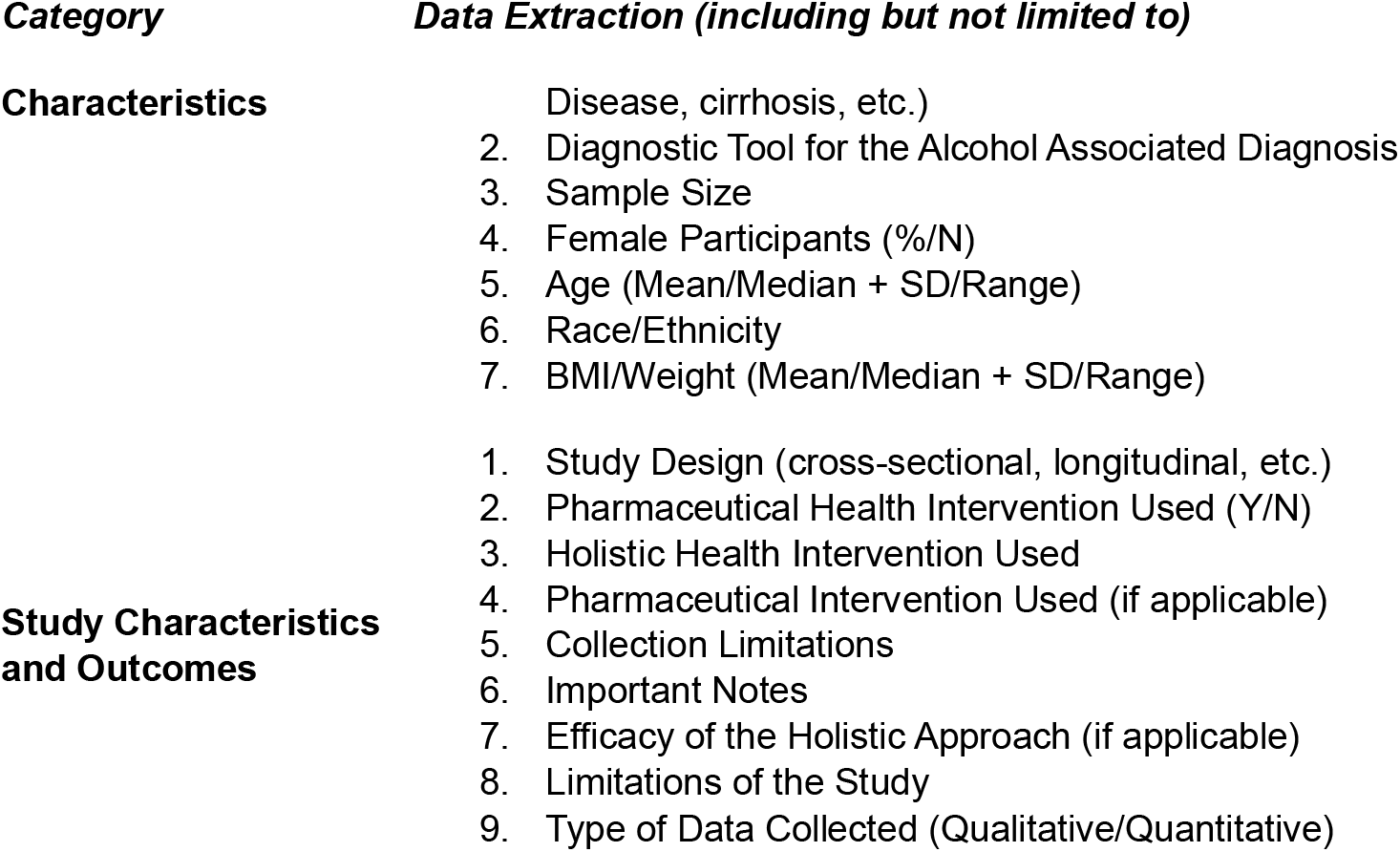
Data Extraction Form.

The following objectives will be considered and reported on:

- Which holistic approaches have been applied to the treatment or management of AUD in the literature?
- What outcome measures were employed to gauge the efficacy of these therapies, and how were they evaluated?
- What justification, if any, is given for the selection of holistic therapies for the treatment of AUD?
- Does the selection of outcome measures used to assess these interventions have any justification?

Additionally, to understand any gaps in the literature, members of the review team will separately tabulate and report on participant characteristics for all studies included in the review as part of the data collection. This will be based on factors such as demographics and social context, to provide insight into the representation of different populations in the studies.

The following queries will be discussed in this review of the data extracted:

- What details on the participants are included in the studies?
- Are socially stratifying elements (such as gender, race, and socioeconomic class) considered and reported?
- What justification, if any, is given for the selection of the study participant groups?

The focus of this review is on identifying the holistic interventions that have been used in AUD treatment, the outcome measures employed, and the representation of different participant characteristics in these studies. The results will be summarized in text, presented in tables for clarity, and published as Appendices. A narrative summary of the identified interventions, outcome measures, and participant characteristics will be provided, including justifications for the choices made in the studies and any information that was not reported. Any implications of these findings will be discussed in the Discussion section of the report. A quality appraisal of the included studies will not be conducted as this is a scoping review designed to assess the current literature, not the quality.

## Ethics and dissemination

This scoping review does not require an ethical review or approval since it is collecting and reviewing material that is publicly available. The review will be submitted for publication and is registered in Open Science Forum. The findings of this review will help other researchers identify and establish current gaps in the usage of holistic interventions for the AUD population.

## Patient/ Public involvement

The research team will not need patient or public involvement for the proposed scoping review.

## Conclusion

Our protocol for a scoping review to synthesize the current literature focused on the use of holistic practices in the treatment of AUD is presented. The scoping review will comprehensively analyze existing research on holistic interventions, such as mind-body therapies, nutritional strategies, and psychosocial support, in the context of AUD treatment, evaluate how these approaches are integrated with conventional care, and identify gaps in the literature to guide future research. Exploring these interventions is critical, as traditional pharmaceutical treatments often fail to address the underlying contributors to AUD, including trauma, mental health conditions, and lifestyle factors. Therefore, a critical next step in addressing this issue is synthesizing currently available literature to understand the state of the science and to stimulate future hypotheses regarding the efficacy and implementation of holistic strategies in AUD recovery. Gaining a deeper understanding of how holistic health practices impact recovery outcomes may inform more comprehensive, person-centered treatment approaches and support sustained abstinence and well-being in individuals affected by AUD.

## Supporting information

Supplemental Materials for Protocol

## Data Availability

Not applicable

https://osf.io/vw7mz

## Abbreviations

AUD: Alcohol Use Disorder
ALD: Alcohol Liver Disease
PRISMA-P: Preferred Reporting Items for Systematic Review and Meta-Analysis Protocols
PRISMA-ScR: Preferred Reporting Items for Systematic Reviews and Meta-Analyses Extension for Scoping Reviews

## Ethics statements

### Patient and public involvement

None to report

### Patient consent for publication

Not applicable.

### Author Statement

SN and JJB conceived the idea and development the research question. SN and AR wrote the initial review draft and developed the study methods. JJB finalized the methods and all authors (SN, AR, GB, DB, GRW, JJB) reviewed and approved the manuscript. The views, information or content, and conclusions presented do not necessarily represent the official position or policy of, nor should any official endorsement be inferred on the part of, the Clinical Center, the National Institutes of Health, or the Department of Health and Human Services.

### Data Statement

Not applicable

## References

1. Association AP. Substance-Related and Addictive Disorders. In Diagnostic and statistical manual of mental disorders. 2022.

2. Grant BF, Goldstein RB, Saha TD, Chou SP, Jung J, Zhang H, et al. Epidemiology of DSM-5 Alcohol Use Disorder: Results From the National Epidemiologic Survey on Alcohol and Related Conditions III. JAMA Psychiatry. 2015;72(8):757–66.

3. Arakelyan S, Mikula-Noble N, Ho L, Lone N, Anand A, Lyall MJ, et al. Effectiveness of holistic assessment-based interventions for adults with multiple long-term conditions and frailty: an umbrella review of systematic reviews. Lancet Healthy Longev. 2023;4(11):e629–e44.

4. Ayares G, Idalsoaga F, Diaz LA, Arnold J, Arab JP. Current Medical Treatment for Alcohol-Associated Liver Disease. J Clin Exp Hepatol. 2022;12(5):1333–48.

5. Zheng S, Edney SM, Goh CH, Tai BC, Mair JL, Castro O, et al. Effectiveness of holistic mobile health interventions on diet, and physical, and mental health outcomes: a systematic review and meta-analysis. EClinicalMedicine. 2023;66:102309.

6. Coriale G, Fiorentino D, De Rosa F, Solombrino S, Scalese B, Ciccarelli R, et al. Treatment of alcohol use disorder from a psychological point of view. Riv Psichiatr. 2018;53(3):141–8.

7. Rosato V, Abenavoli L, Federico A, Masarone M, Persico M. Pharmacotherapy of alcoholic liver disease in clinical practice. Int J Clin Pract. 2016;70(2):119–31.

8. Fuller RK, Hiller-Sturmhofel S. Alcoholism treatment in the United States. An overview. Alcohol Res Health. 1999;23(2):69–77.

9. Kelly JF, Humphreys K, Ferri M. Alcoholics Anonymous and other 12-step programs for alcohol use disorder. Cochrane Database Syst Rev. 2020;3(3):CD012880.

10. Perez-Reytor D, Karahanian E. Alcohol use disorder, neuroinflammation, and intake of dietary fibers: a new approach for treatment. Am J Drug Alcohol Abuse. 2023;49(3):283–9.

11. Morris J, Boness CL, Burton R. (Mis)understanding alcohol use disorder: Making the case for a public health first approach. Drug Alcohol Depend. 2023;253:111019.

12. Singal AK, Charlton MR. Nutrition in alcoholic liver disease. Clin Liver Dis. 2012;16(4):805–26.

13. Arksey HOM, Lisa. Scoping studies: towards a methodological framework. International Journal of Social Research Methodology. 2005;8(1):19–32.

14. Peters MDJ GC, McInerney P, Munn Z, Tricco AC, Khalil, H. Aromataris E, Lockwood C, Porritt K, Pilla B, Jordan Z, editors.. Scoping Reviews. JBI Manual for Evidence Synthesis. 2020.

15. Tricco AC, Lillie E, Zarin W, O’Brien KK, Colquhoun H, Levac D, et al. PRISMA Extension for Scoping Reviews (PRISMA-ScR): Checklist and Explanation. Ann Intern Med. 2018;169(7):467–73.

16. Levac D, Colquhoun H, O’Brien KK. Scoping studies: advancing the methodology. Implement Sci. 2010;5.

17. Micah DJ Peters CG, Patricia McInerney, Zachary Munn, Andrea C. Tricco, Hanan Khalil. Scoping Reviews. JBI Manual for Evidence Synthesis. 2024.

18. Covidence. Covidence systematic review software, Veritas Health Innovation, Melbourne, Australia. 2023.

