## Supplemental Materials for Protocol for "Assessing Holistic Health Interventions and Approaches for Individuals with Alcohol Use Disorder (AUD): Protocol for a Scoping Review"

Supplemental Table 1: MeSH Terms Used for Search Strategy in all Databases

| Concept | Search Strategy |
| --- | --- |
| AUD | "Alcohol-Related Disorders"[Mesh:NoExp] OR "Alcohol Drinking"[Mesh] OR "Alcoholism"[Mesh] OR "Alcoholic Intoxication"[Mesh] OR "alcoholism"[Title/Abstract] OR "alcohol drink*"[Title/Abstract] OR "alcohol use"[Title/Abstract] OR "alcohol abuse"[Title/Abstract] OR "alcohol habit*"[Title/Abstract] OR "alcohol misus*"[Title/Abstract] OR "alcohol addiction"[Title/Abstract] OR "alcohol consumption*"[Title/Abstract] OR "alcohol intake*"[Title/Abstract] OR "binge drinking"[Title/Abstract] OR "alcohol related"[Title/Abstract] OR "alcohol problem*"[Title/Abstract] OR "alcoholic*"[Title/Abstract] OR "alcohol* intoxicat*"[Title/Abstract] OR "alcohol depend*"[Title/Abstract] OR "alcohol deterrent*”[Title/Abstract] OR "antialcoholic*"[Title/Abstract] OR "alcohol withdrawal*"[Title/Abstract] OR "alcohol abstinence*"[Title/Abstract] OR "alcohol disorder*"[Title/Abstract] |
| Holistic | "Complementary Therapies"[Mesh] OR "complimentary medicine*"[Title/Abstract] OR "holistic health*"[Title/Abstract] OR "holistic healing"[Title/Abstract] OR "holistic intervention*"[Title/Abstract] OR "homeopath*"[Title/Abstract] OR "naturopath*"[Title/Abstract] OR "alternative medicine*"[Title/Abstract] OR "integrat* medicine*"[Title/Abstract] OR "alternative therap*"[Title/Abstract] OR "ayurv*"[Title/Abstract] OR "eastern medicine*"[Title/Abstract] OR "Chinese medicine*"[Title/Abstract] OR "Acupuncture Therapy"[Mesh] OR "acupuncture*"[Title/Abstract] OR "music therap*"[Title/Abstract] OR "art therap*"[Title/Abstract] OR "relaxation therap*"[Title/Abstract] OR "relaxation treatment*"[Title/Abstract] OR "Sleep Quality"[Mesh] OR "sleep quality"[Title/Abstract] OR "guided imagery"[Title/Abstract] OR "biofeedback*"[Title/Abstract] OR "stretching exercis*"[Title/Abstract] OR "exercise treatment*"[Title/Abstract] OR "exercise intervention*"[Title/Abstract] OR "exercise therap*"[Title/Abstract] OR "buteyko"[Title/Abstract] OR "breathing exercis*"[Title/Abstract] OR "meditation*"[Title/Abstract] OR "mindfulness"[Title/Abstract] OR "mind body"[Title/Abstract] OR "horticultural*"[Title/Abstract] OR "hypnosis"[Title/Abstract] OR "Exercise Movement Techniques"[Mesh] OR "yoga"[Title/Abstract] OR "Reiki"[Title/Abstract] OR "tai chi"[Title/Abstract] OR "Pilates"[Title/Abstract] OR "Massage"[Mesh] OR "massage*"[Title/Abstract] OR "faith healing"[Title/Abstract] OR "spiritual healing"[Title/Abstract] OR "herbal remed*"[Title/Abstract] OR "herbal therap*"[Title/Abstract] OR "Thiamine"[Mesh] OR "thiamine"[Title/Abstract] OR "Diet Therapy"[Mesh:NoExp] OR "diet* therap*"[Title/Abstract] OR "diet* treatment*"[Title/Abstract] OR "diet* restrict*"[Title/Abstract] OR "diet* modif*"[Title/Abstract] OR "Diet, Ketogenic"[Mesh] OR "ketogenic diet*"[Title/Abstract] OR "whole food diet*"[Title/Abstract] OR "nutrient dense*"[Title/Abstract] OR "Dietary Supplements"[Mesh] OR "diet* supplement*"[Title/Abstract] OR "vitamin* supplement*"[Title/Abstract] OR "herb* supplement*"[Title/Abstract] OR "food supplement*"[Title/Abstract] OR "probiotic*"[Title/Abstract] OR "prebiotic*"[Title/Abstract] OR "Self-Help Groups"[Mesh] OR "self-help group*"[Title/Abstract] OR "support group*"[Title/Abstract] OR "Community Support"[Mesh] OR "community support*"[Title/Abstract] |
| Intervention | "Clinical Trials as Topic"[Mesh] OR "Clinical Trial" [Publication Type] OR "clinical trial*"[Title/Abstract] OR "clinical stud*"[Title/Abstract] OR "controlled clinical"[Title/Abstract] OR "Random Allocation"[Mesh] OR "random allocat*"[Title/Abstract] OR "randomized"[Title/Abstract] OR "randomly"[Title/Abstract] OR "trial*"[Title] OR "Cohort Studies"[Mesh] OR "cohort stud*"[Title/Abstract] OR "followup stud*"[Title/Abstract] OR "follow up stud*"[Title/Abstract] OR "longitudinal"[Title/Abstract] OR "retrospective"[Title/Abstract] OR "prospective"[Title/Abstract] OR "Observational Study" [Publication Type] OR "Observational Studies as Topic"[Mesh] OR "observational study"[Title/Abstract:~2] OR "observational studies"[Title/Abstract:~2] OR "Qualitative Research"[Mesh] OR "qualitative research"[Title/Abstract] OR "qualitative stud*"[Title/Abstract] |
| Combined Concepts and Limits | (((#1 AND #2 AND #3) NOT ("Animals"[Mesh] NOT ("Animals"[Mesh] AND "Humans"[Mesh]))) NOT ("Models, Animal"[Mesh] OR "In Vitro Techniques"[Mesh] OR "mice"[Title/Abstract] OR "mouse"[Title/Abstract] OR "rats"[Title] OR "Rodentia"[Mesh] OR "rodent*"[Title/Abstract] OR "pig"[Title/Abstract] OR "pigs"[Title/Abstract] OR "Dogs"[Mesh] OR "dog"[Title/Abstract] OR "dogs"[Title/Abstract])) NOT ("Letter"[Publication Type] OR "Editorial" [Publication Type] OR "Comment"[Publication Type] OR "News"[Publication Type] OR "Retracted Publication"[Publication Type] OR "Retraction of Publication"[Publication Type] OR "retraction of publication"[Title/Abstract] OR "retraction notice"[Title] OR "retracted publication"[Title/Abstract] OR "protocol*"[Title] OR "symposium*"[Title/Abstract] OR "Congress"[Publication Type] OR "Consensus Development Conference"[Publication Type] OR "conference abstract*"[Title/Abstract] OR "conference proceeding*"[Title/Abstract] OR "conference paper*"[Title/Abstract] OR "conference review*"[Title/Abstract]) Filters: English |

­­
